# The cost of illness for childhood clinical pneumonia and invasive pneumococcal disease in Nigeria

**DOI:** 10.1101/2021.08.02.21261497

**Authors:** Aishatu A Adamu, Boniface Karia, Musa M Bello, Mahmud G Jahun, Safiya Gambo, J Ojal, J. Anthony G Scott, J Jemutai, Ifedayo M.O. Adetifa

## Abstract

**Background:** Pneumococcal disease contributes significantly to childhood morbidity and mortality and treatment is costly. Nigeria recently introduced the Pneumococcal Conjugate Vaccine (PCV) to prevent pneumococcal disease. The aim of this study is to estimate health provider and household costs for the treatment of pneumococcal disease in children aged <5 years (U5s), and to assess the impact of these costs on household income.

**Methods:** We recruited U5s with clinical pneumonia, pneumococcal meningitis or pneumococcal septicaemia from a tertiary and a secondary level hospital in Kano, Nigeria. We obtained resource utilisation data from medical records to estimate costs of treatment to provider, and household expenses and income loss data from caregiver interviews to estimate costs of treatment to households. We defined catastrophic health expenditure (CHE) as household costs exceeding 25% of monthly household income and estimated the proportion of households that experienced it. We compared CHE across tertiles of household income (from the poorest to least poor).

**Results:** Of 480 participants recruited, 244 had outpatient pneumonia, and 236 were hospitalised with pneumonia (117), septicaemia (66) and meningitis (53). Median (IQR) provider costs were US$17 (US$14-22) for outpatients and US$272 (US$271-360) for inpatients. Median household cost was US$51 (US$40-69). Overall, 33% of households experienced CHE, while 53% and 4% of the poorest and least poor households, experienced CHE respectively. The odds of CHE increased with admission at the secondary hospital, a diagnosis of meningitis or septicaemia, higher provider costs, and caregiver having a non-salaried job.

**Conclusion:** Provider costs are substantial, and households incur treatment expenses that considerably impact on their income and this is particularly so for the poorest households. Sustaining the PCV programme and ensuring high and equitable coverage to lower disease burden will reduce the economic burden of pneumococcal disease to the healthcare provider and households.

What is already known?

- Children <5 years have the highest incidence of pneumonia and invasive pneumococcal disease (IPD) and Nigeria bears the largest burden in sub-Saharan Africa.
- Pneumococcal conjugate vaccine (PCV) was introduced in Nigeria in 2016 to reduce the burden of pneumococcal disease
- PCV is currently subsidised through Gavi financial support and Nigeria will transition to full self-financing in a few of years
- There is no contextual evidence in Nigeria on economic burden of IPD to the health system and society that can support longer term investments in PCV when Gavi co-financing terminates

What are the new findings?

- Treatment of one hospitalised episode of pneumococcal disease cost on average, US$300 to the provider, and US$83 to the household with significant variation by clinical syndrome and level of care
- Overall, a third of the households encountered costs that were catastrophic (i.e., >25% of household income)
- Burden of CHE varied by household income tertile ranging from 4% in the least poor households (highest income tertile) to 53% in the poorest households (lowest income tertile)
- Despite the short illness duration, pneumococcal disease syndromes result in huge economic costs to providers and households

What do the new findings imply?

- Sustaining the PCV programme and achieving high PCV coverage has the potential of saving resources at both provider and household level
- Households are at risk of further impoverishment from catastrophic expenses associated with treatment of pneumococcal disease. This risk can also be significantly reduced by PCV.

## INTRODUCTION

Introduction of the Pneumococcal Conjugate Vaccine (PCV) has significantly reduced the global burden of pneumococcal disease.[1] Despite availability of effective vaccination, pneumococcal disease syndromes remain leading causes of preventable morbidity, mortality and economic burden, particularly among children aged <5 years (U5s) and in low- and middle-income countries (LMICs)).[1,2] In 2015, there were still ∼ 9 million cases of IPD in U5 children resulting in over 300,000 deaths despite this being a significant decline of >60% from the pre-vaccination PCV era.[1,3] Slow uptake and sub-optimal coverage of PCV are partly responsible for a disproportionate pneumococcal disease burden in LMICs in the post-PCV era.[1] Unsurprisingly, pneumococcal diseases are associated with substantial annual economic health system costs of about US$13.7 billion and societal costs of US$14.3 billion globally.[4] Although associated with substantial vaccine and delivery costs, ranging between US$52 in Africa to US$599 in Europe per vaccinated child, the introduction of PCVs to infant immunisation programmes is expected to provide savings estimated as US$3.2 billion from averted hospital visits and care, and an additional US$2.6 billion from societal costs globally.[4] Economic cost studies on pneumococcal diseases report substantial costs of treatment to healthcare provider, households and families, with significant out-of-pocket (OOP) payment for health particularly in low-and middle-income countries (LMICs).[5–12] OOP payment for health care can result in catastrophic expenses capable of driving households further into poverty.

With >1 million pneumococcal disease cases resulting in nearly 50,000 deaths among U5s in 2015, Nigeria has the highest burden of pneumococcal disease in sub-Saharan Africa (sSA).[1] Approximately 40% of Nigeria’s population live below the poverty line, and 15% of the population incur healthcare expenses from an illness episode that exceeds 10% of their household income annually. [13,14] In addition, 2.3% are pushed into poverty by these health expenses.[14]

Financing of healthcare in Nigeria is via multiple and largely uncoordinated channels.[15] It has one of the lowest health insurance coverage in sSA because the National Health Insurance Scheme (NHIS) currently targets persons employed in the formal sector, which represent about 5% of the population.[16,17] The huge informal sector largely finances health care through OOP payment that are over three-quarters of total expenditure on health.[18] The consequences of the huge pneumococcal disease burden and limited financial protection, especially for the poor, extend beyond the clinical as households are at high risk of impoverishment. Additionally, to avoid such unexpected financial burden, households can delay or refrain from seeking healthcare and this ultimately results in greater costs and/or poorer outcomes.[21] Existing mechanisms to provide financial protection to households range from subsidised services for vulnerable populations such as U5s and pregnant women, to the recent expansion of community-based health insurance (CBHI) to the informal sector.[19,20]

Ahead of Nigeria completing the transition to full self-financing of PCV in 2028,[22] data on the economic burden of pneumococcal diseases will help inform the policy required to assure sustainability of the PCV programme. A current description of the costs of treating childhood pneumococcal diseases in Nigeria is lacking highlighting a significant data gap.

The objectives of this study are to: 1) to estimate the provider costs of outpatient and inpatient clinical pneumonia, and inpatient pneumococcal septicaemia and meningitis 2) to estimate the household costs of hospitalised clinical pneumonia, and pneumococcal meningitis and bacteraemia; and 3) to assess the economic impact to households of hospitalisation with clinical pneumonia, and pneumococcal septicaemia and meningitis among U5s in Kano, northern Nigeria.

## METHODS

### Study design and setting

This was a cross sectional study conducted at the two largest paediatrics units in Kano, Kano State in northern Nigeria - Aminu Kano Teaching Hospital (AKTH) and Murtala Mohamed Specialist Hospital (MMSH) which serve an overlapping catchment population. Kano is the most densely populated city in the region with an estimated population of 12.2million (∼1.3million U5s) occupying 20,760km^2^.[23,24] About 55% of the population in Kano state reside in households below the poverty line.[13] The infant and U5 mortality per 1,000 live births in Kano (National) were 112 (70) and 203 (120) in 2018.[25]

### Study population

Children were recruited prospectively and were eligible if aged 1-59 months, presented to AKTH or MMSH and had at least one of three possible diagnoses of interest. These were (i) clinical pneumonia (ii) pneumococcal septicaemia (iii) pneumococcal meningitis. We excluded children that died during admission.

Sample sizes of 100, 50 and 30 were expected to provide a cost estimate for outpatient clinical pneumonia, inpatient pneumonia, and septicaemia and meningitis based on standard deviation (±precision) of US$5 (±US$1), US$21 (±US$6) and US$33 ± US$12) respectively.[8,9,26].

### Data collection

We recruited outpatient pneumonia cases and interviewed caregivers on the day of diagnosis. For inpatient pneumonia cases, eligible children admitted 8am to 4pm were recruited on the day of admission; those admitted 4pm to 8am the next morning were enrolled on the next day. For septicaemia and meningitis cases, participants were recruited when confirmatory laboratory results were available. We collected data between January and October 2020. For each hospital, we recruited a volunteer nurse not directly involved in clinical care to collect data. We used structured quantitative tools adapted from a similar study in The Gambia for data collection.[9]

We extracted resource use data such as length of hospital stay, type and quantity of medications and intravenous fluids used, laboratory investigations and other specialised services including blood transfusion and use of oxygen from patients’ records, case folders, prescriptions, and laboratory request forms. We obtained unit costs of hospital resources e.g., medication, fluids, etc utilised from the respective hospitals (see Table S1).

Sociodemographic characteristics, OOP costs, non-medical expenses, productivity time loss, household income and sources of finances used to pay the treatment costs were obtained through caregiver interviews. Additional data on household income and sources of finances used to pay the treatment costs were also collected.

### Cost components

We collected provider costs, and direct and indirect household costs. Provider costs

Provider costs included costs of direct healthcare services i.e., costs of medications, laboratory investigations, intravenous fluids, oxygen, blood transfusion and inpatient bed-day. We used full costs for drugs and only applied dose-specific costs if the drug was re-useable and the residuals amounted to another full dose. For instance, for a re-useable drug, if a unit dose was 1,000mg and 500mg was administered, the cost per dose would be half of the unit cost. We obtained cost of oxygen from the nursing unit, and for blood transfusion, we used previously published costs.[27]

The inpatient bed-day is the daily stay cost or the ‘hotel’ component and comprises costs of food, personnel, and utilities. We used the average 2020 admission charge for AKTH and assumed admission costs at MMSH to be 60% of AKTH since admission is ‘free’ to patients at MMSH. Studies have found up to 60-70% differences in bed-day costs between tertiary and secondary-level hospitals.[8,9]

#### Household costs

We collected direct healthcare costs to households which are user fees related to consultations, investigations, and medications incurred from date of admission to date of discharge. Non-healthcare costs were the costs of transportation, accommodation and feeding incurred during admission (from date of admission to date of discharge) by main and accompanying caregiver. The accompanying caregiver was defined as any household member that assisted the main caregiver with care of the patient during the admission.

Preadmission costs data were collected either on the day of recruitment or on the earliest convenient day for the caregiver, while data on costs incurred over the course of admission were obtained at or close to discharge.

We collected data on caregiver’s income and productivity time loss due to time away from their usual activities owing to illness.

### Data analysis

Data were analysed using STATA 15.1 (Stata Corp LP, College Station, Texas). All costs were converted to US$ using average 2020 conversion rates 1 US$ = 360.5 NGN (Central bank of Nigeria).[28]

We summed up the components of respective cost categories for provider costs and direct household costs. We estimated indirect costs using the human-capital approach (HCA) by estimating income lost by caregiver(s) due to absence from work per day spent caring for the child. Self-employed caregivers were asked to give an estimate of daily earnings while those on monthly salary were asked to state their monthly wage from which daily income was calculated. Indirect costs were then calculated as daily income multiplied by the number of days taken off from work.

We present costs from the health provider and household’s perspectives along with their components separately as means with standard deviation (SD), and medians with interquartile range (IQR). We used Kruskal-Wallis test to assess differences in costs between disease categories and Wilcoxon rank sum test to compare costs between the two hospitals.

We evaluated the impact of health expenditures on available household resources by assessing direct, indirect, and total costs as respective proportions of household income. We used household income to categorise households into tertiles from the poorest (tertile 1) to least poor (tertile 3). We used Kruskal-Wallis tests to compare costs as a proportion of household income across household income tertiles.[29] We also evaluated catastrophic health expenditure (CHE) as costs exceeding a specified threshold of household available resources.[30] In this analysis, we used household income as a measure of available resources and set the base threshold as 25%.[14,31,32] We defined costs as catastrophic if they exceeded 25% of household monthly income (CHE_25_) and also explored impact at 10% (CHE_10_) and 40% (CHE_40_) thresholds. We used multivariable logistic regression models to identify factors associated with CHE_25_. Independent variables that were associated with CHE_25_ at significance level p=0.1 were sequentially added to the model and kept if they were significantly associated with cost (p<0.05) or changed effects of included variables.

Excluded variables were then re-introduced to check if they further changed the effect sizes of included variables. Adjusted odds ratios and P values from the likelihood ratio test (LRT) are reported.

### Sensitivity analysis

We conducted one-way sensitivity analyses of provider costs by varying the source of bed-day costs. We used the average cost per inpatient bed-day for tertiary and secondary facilities in Nigeria from the WHO-CHOICE after accounting for inflation.[33] We also conducted a sensitivity analysis of indirect costs by using the Willingness to pay (WTP) approach to assess productivity loss. Indirect costs using WTP approach were calculated as the product of the amount caregivers were willing to pay for main activity they would have been otherwise engaged in and the total days taken off from work due to childcare.

### Patient involvement

Patients were not directly involved in the design, conduct, reporting or dissemination plans of this research.

## RESULTS

### Study participants

On average about 97% of eligible participants were enrolled. A total of 480 children (244 outpatient pneumonia, 117 inpatient pneumonia, 53 meningitis and 66 septicaemia) were enrolled (see table S2). Of these, 387, (81%) were aged ≥1 year. Clinical pneumonia cases were younger than their counterparts with meningitis or septicaemia (mean age, 19 vs. 25 months, p=0.002). Caregivers were aged 20 to 48 years, mostly mothers, had at least secondary school level education and were employed. Caregivers of children with outpatient pneumonia were more likely to be unemployed compared to those with hospitalised children (20% vs 3%, p= <0.0001). The mean duration of hospitalisation was five days for cases with pneumonia or septicaemia but seven days for those with meningitis. Majority (362/480, 75.4%) of the children had sought care prior to index visit/hospitalisation.

### Provider costs for outpatient pneumonia

The median provider cost for outpatient pneumonia was US$17 (IQR:14-22), and was higher in AKTH (US$20, IQR:14-23) compared to MMSH (US$16; IQR:14-19, p=0.0002). Overall, costs for outpatient clinic visit, medications and investigations accounted for 43%, 37% and 20% of provider costs. The median costs for laboratory investigations were higher in AKTH US$ 7 (IQR:0-8) compared to MMSH US$0 (IQR:0-4, p<0.0001). Medications costs were similar between the two hospitals. Median expenses on seeking care elsewhere prior to index presentation were US$9 (IQR:0-13) in AKTH and US$8 (IQR:5-13) in MMSH (p=0.70).

### Provider costs for hospitalised children

The respective median/mean provider costs, as shown in table 1, were highest for meningitis in both hospitals which was mostly driven by bed-day costs. The median provider costs (all syndromes combined) were significantly higher in AKTH (US$359, IQR:308-400) compared to MMSH (US$223, IQR:196-264, p<0.0001).

**Table 1:**
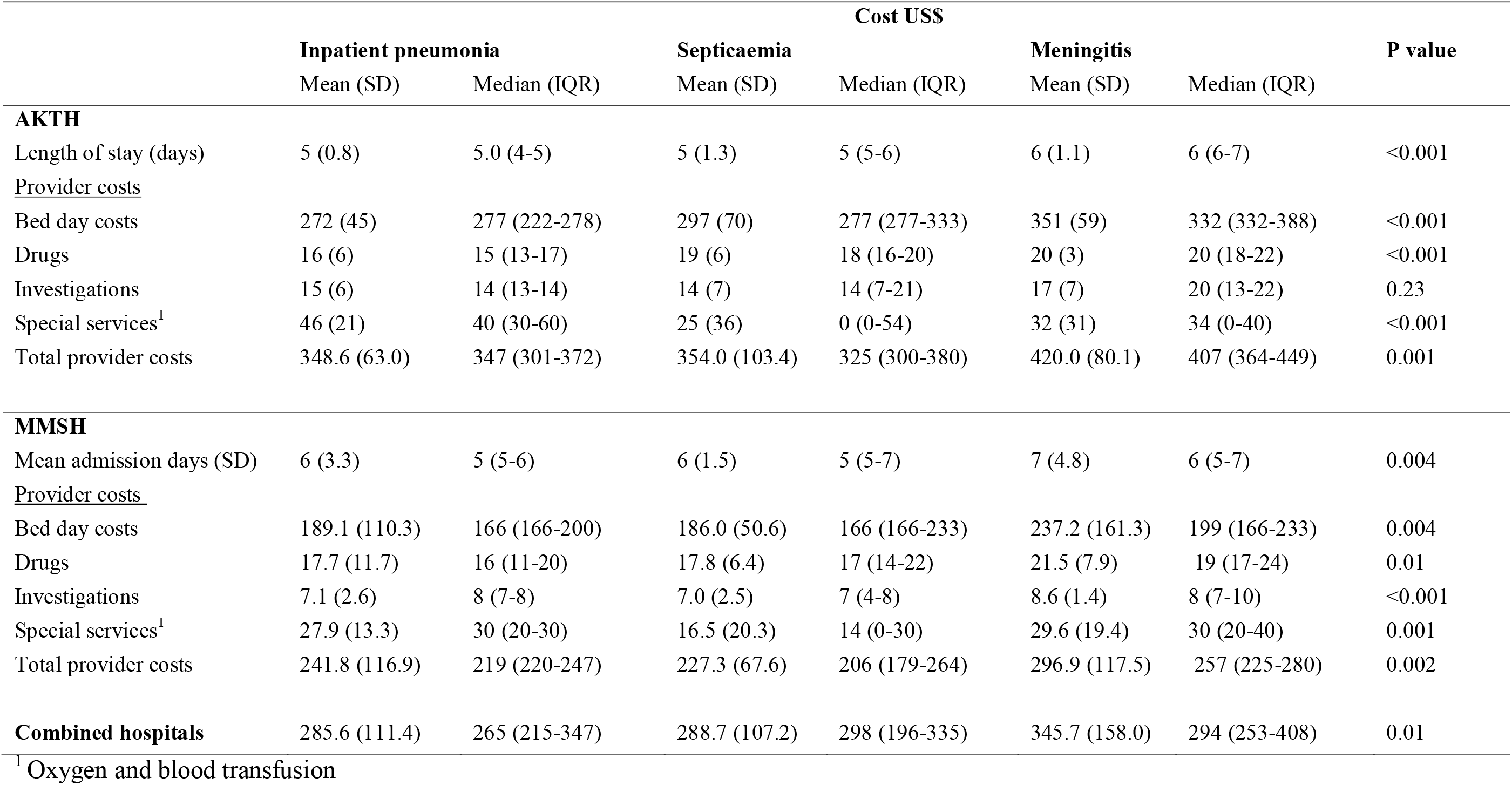
Provider costs for inpatient pneumonia, meningitis, and septicaemia in US$.

### Household costs for hospitalised children

Median household income was similar between disease categories but was significantly higher for those presenting to AKTH (US$250, IQR:222-277, p=0.02) compared to MMSH (US$222, IQR:194-277). Majority of caregivers (217/236, 92%) reported using a combination of current income and savings to cover expenses. Only about 3% reported using other sources such as borrowing, asking relatives, or selling assets to cover expenses.

Median direct household costs as shown in table 2 were highest for meningitis and lowest for pneumonia in both hospitals. However, there was no significant difference in overall direct household costs between the two hospitals. Direct costs comprised mostly of user fees, and for each disease category in both hospitals, medication costs were the largest fraction of user fees. (Figure S1A-B)

**Table 2:** Direct and indirect costs to households per illness episode (US$)

| | Cost US\$ | | | | | | |
| --- | --- | --- | --- | --- | --- | --- | --- |
|  | Inpatient Pneumonia |  | Septicaemia |  | Meningitis |  | P value |
|  | Mean (SD) | Median (IQR) | Mean (SD) | Median (IQR) | Mean (SD) | Median (IQR) |  |
| AKTH |  |  |  |  |  |  |  |
| Pre-admission costs | 9.4 (9.3) | 8.6 (0-12) | 19.8 (26.4) | 13.9 (8-26) | 14.9 (11.0) | 13.6 (8-25) | 0.01 |
| Current admission costs |  |  |  |  |  |  |  |
| 1. <u>Direct costs</u> |  |  |  |  |  |  |  |
| Transportation | 1.1 (1.9) | 0.0 (0-4) | 2.1 (5.7) | 0.0 (0-5) | 1.1 (2.9) | 0.0 (0-0) | 0.84 |
| Feeding | 0.6 (2.2) | 0.0 (0-0) | 1.2 (4.8) | 0.0 (0-0) | 1.3 (6.1) | 0.0 (0-0) | 0.98 |
| User fees | 38.5 (29.3) | 35.2 (30-38) | 44.5 (25.4) | 35.8 (32-47) | 44.3 (6.7) | 44.3 (39-50) | <0.001 |
| Total direct household costs | 50 (30.0) | 43.6 (36-55) | 73.5 (61.1) | 60.7 (45-75) | 64.3 (16.8) | 68.8 (51-76) | <0.001 |
| 2. <u>Indirect costs</u> |  |  |  |  |  |  |  |
| Working days lost | 5 (0.9) | 5 (4-5) | 5 (1.2) | 5 (5-6) | 6 (1.1) | 6 (6-7) | <0.001 |
| Daily income lost | 4 (2) | 4 (3-5) | 4 (2) | 5 (3-5) | 5 (2) | 5 (4-5) | 0.23 |
| Total indirect costs | 19 (7.) | 19 (14-23) | 22 (12.0) | 23 (15-23) | 28 (14.7) | 27.7 (22-28) | 0.006 |
| Total household costs | 68.9 (28.2) | 64.5 (55-75) | 95.4 (63.8) | 82.6 (63-96) | 92.3 (22.1) | 89.3 (79-106) | <0.001 |
| MMSH |  |  |  |  |  |  |  |
| Working days lost |  |  |  |  |  |  |  |
| Pre-admission costs | 12.0 (9.8) | 9.9 (7-12) | 12.2 (10.4) | 11.4 (0-14) | 18.6 (10.4) | 17.7 (11-26) | 0.003 |
| Current admission costs |  |  |  |  |  |  |  |
| 1. <u>Direct costs</u> |  |  |  |  |  |  |  |
| Transportation | 2.9 (4.8) | 1.9 (0-3) | 1.7 (3.9) | 0.0 (0-5) | 2.9 (4.4) | 0.0 (0-6) | 0.17 |
| Feeding | 1.1 (3.4) | 0.0 (0-0) | 3.2 (12.8) | 0.0 (0-0) | 6.8 (17.8) | 0.0 (0-0) | 0.57 |
| User fees | 27.8 (11.3) | 29.1 (22-36) | 50.7 (84.3) | 34.4 (29-44) | 59.2 (94.3) | 40.3 (36-45) | <0.001 |
|  | Inpatient Pneumonia |  | Septicaemia |  | Meningitis |  | P value |
|  | Mean (SD) | Median (IQR) | Mean (SD) | Median (IQR) | Mean (SD) | Median (IQR) |  |
| Total direct household costs | 43.7 (14.4) | 42.7 (35-53) | 74.3 (102.0) | 53.8 (42-72) | 92.2 (106.6) | 71.8 (58-83) | <0.001 |
| 2. <u>Indirect cost</u> |  |  |  |  |  |  |  |
| Working days lost | 5.3 (3.3) | 5.0 (4-5) | 5.6 (1.5) | 5.0 (5-7) | 7.4 (6.5) | 6.0 (5-7) | <0.001 |
| Daily income lost | 3 (2) | 3 (2-5) | 4 (2) | 4 (3-5) | 4 (3) | 4 (3-5) |  |
| Total indirect costs | 16.3 (12.4) | 16.6 (7-23) | 21.1 (12.0) | 18.5 (114-26) | 28.4 (22.5) | 22.4 (19-28) | <0.001 |
| Total household costs | 60.0 (21.8) | 59.1 (47-74) | 95.4 (105.7) | 68.4 (60-95) | 120.6 (123.2) | 97.6 (83-108) | <0.001 |
| **Combined hospitals** |  |  |  |  |  |  |  |
| Direct cost | 46.3 (22.3) | 43.6 (36-53) | 73.9 (84.1) | 57.3 (42.2) | 81.2 (84.1) | 69.6 (55-80) | <0.001 |
| Indirect cost | 17.4 (10.6) | 18.5 (9-23) | 21.5 (11.9) | 19.4 (14-26) | 28.2 (19.6) | 23 (19-28) | <0.001 |
| Total cost | 63.7 (24.9) | 62.2 (50-74) | 95.4 (87.3) | 72.2 (61-95) | 109.4 (97.1) | 93.8 (81-106) | <0.001 |

Median indirect costs were lowest for inpatient pneumonia compared to meningitis or septicaemia, as shown in table 2. Comparison between the hospitals, showed indirect costs were slightly higher in AKTH compared to MMSH (US$22 [IQR:15-26] vs. US$18 [IQR:10- 23], p=0.04).

### Economic impact to households

The poorest households spent a median of 25% of their monthly income directly on treatment costs and lost an additional 8% from loss of caregiver time, compared to 13% of income and 6% of caregiver time for the least poor households (data not shown). Treatment costs as fractions of monthly household income were inversely related to household income tertiles (see table S3).

Table 3 shows the proportion of households encountering CHE_25_ at different threshold cut-off values across household income levels. CHE was substantial, and at 10% threshold nearly all households across all income levels encountered CHE. CHE increased with decreasing household income level. This inverse relationship is further illustrated in Fig 1 which shows that as the threshold values increase, the proportions of households encountering catastrophic costs declines steeply for the higher income households, with slowest decline for the poorest households (tertile 1).

**Table 3:** Proportions of households with CHE at different thresholds of household income.

| Income tertile | CHE threshold level |  |  |  |  |  |
| --- | --- | --- | --- | --- | --- | --- |
|  | 10% |  | 25% |  | 40% |  |
|  | % | 95% CI | % | 95% CI | % | 95% CI |
| Tertile 1<br>(poorest) | 97.2 | 92.2 – 99.4 | 53.2 | 43.4 - 62.8 | 18.3 | 11.6 – 26.9 |
| Tertile 2 | 97.2 | 90.3 – 99.7 | 25.0 | 15.5 - 36.6 | 2.8 | 0.3 - 9.7 |
| Tertile 3<br>(least poor) | 81.8 | 69.1 – 90.9 | 3.6 | 0.4 - 12.5 | 0.0 | 0.0 - 0.0 |
| Overall | 93.6 | 89.7 - 96.1 | 33.1 | 27.1 - 39.4 | 9.3 | 5.9 -13.7 |

**Figure 1:**
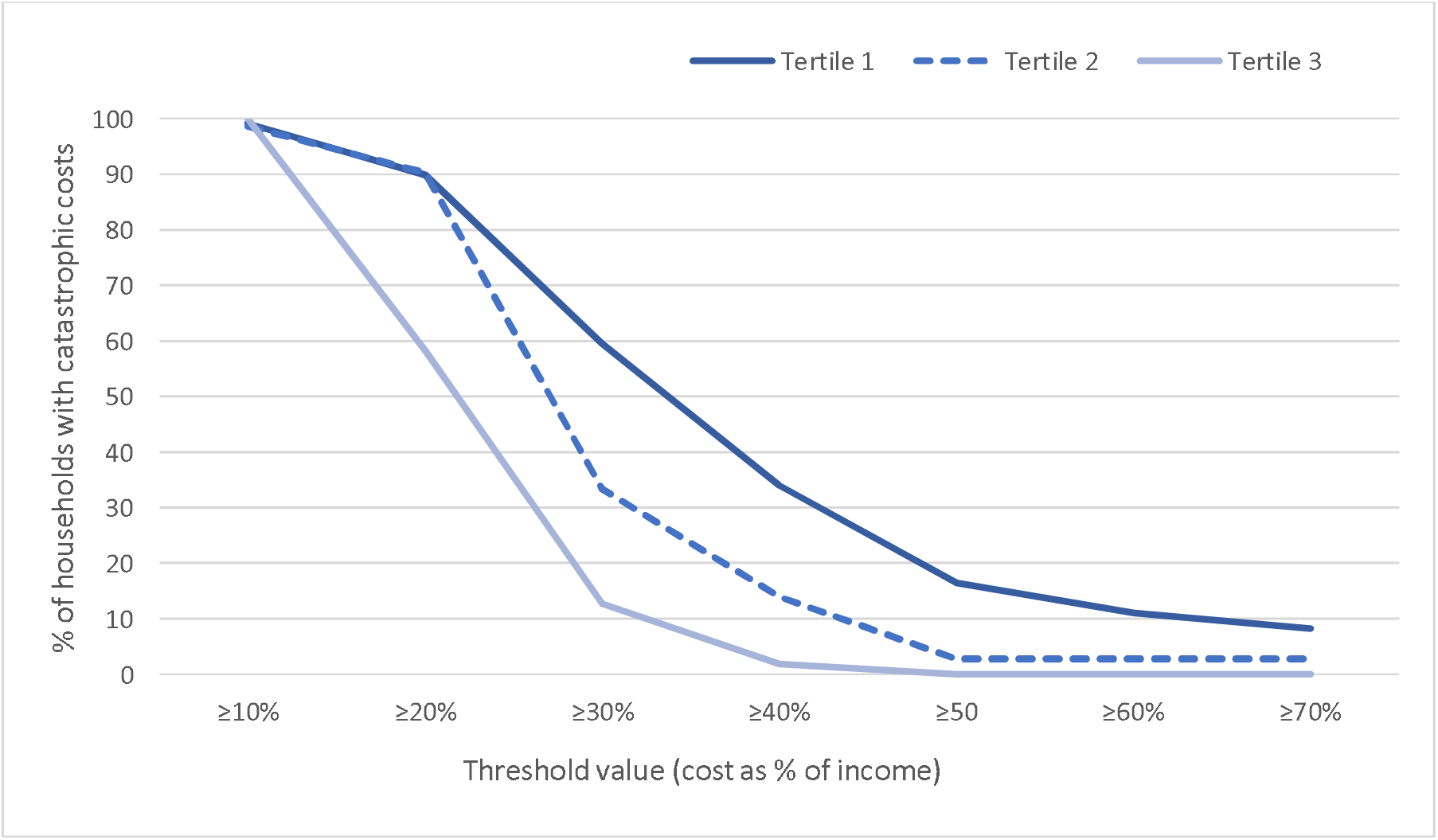
Distribution of catastrophic costs by household income level and threshold value.

Admitting hospital (MMSH), meningitis or septicaemia, seeking care at a private hospital prior to admission and higher provider costs were associated with increased odds of CHE_25_, while having ≥3 U5 children and higher indirect costs lowered the odds of CHE_25_ (see table 4).

**Table 4:**
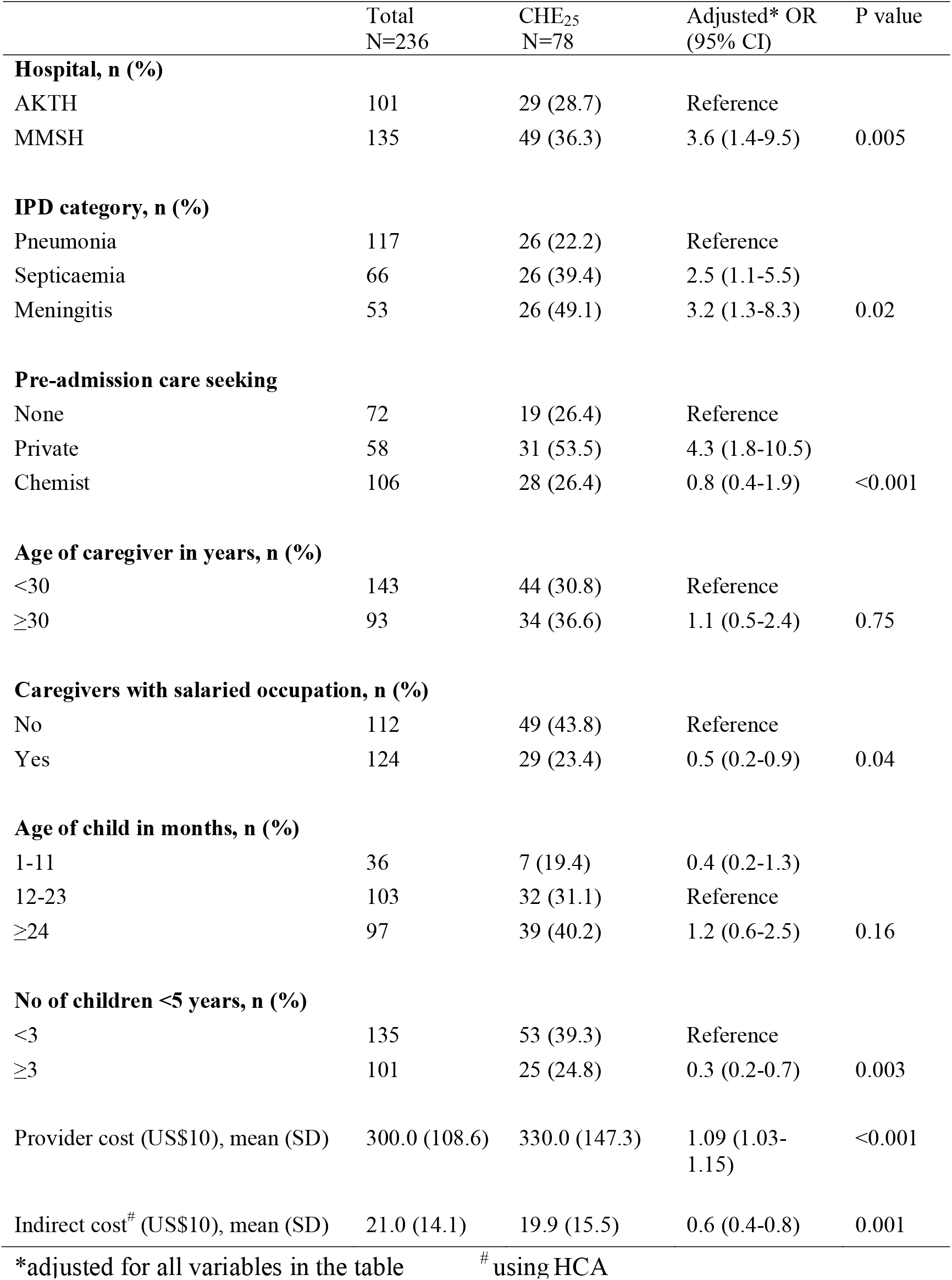
Distribution of CHE_25_ and multivariable logistic regression of factors associated with CHE_25_.

### Sensitivity analyses

Provider costs were sensitive to source of hospital (bed-day) costs. Provider costs were between 38% and 40% lower across all disease categories and between the hospitals when the WHO-CHOICE estimates were used (table S4) compared to actual hospital admission costs (table S4). Indirect costs were also sensitive to approach used as costs were higher for all conditions in both hospitals with the WTP (table S4) compared to the HCA. However, in contrast to HCA, indirect costs were similar between the hospitals (p=0.45)

## DISCUSSION

In this study, we estimated the costs of treatment of clinical pneumonia, septicaemia, and meningitis in Nigerian children as well as the economic impact of these costs on households. Costs varied by hospital as they were higher in the tertiary hospital (AKTH) for all disease categories. Provider costs also varied by illness and were highest for meningitis irrespective of hospital. Costs to households were similar between the hospitals but highest for meningitis and lowest for pneumonia. The economic impact to households was considerable with total costs to households ranging between 25-37% of monthly household income for 5-7 days of hospitalisation. One third of the households incurred CHE at 25% threshold and the poorest households bore the greatest burden of CHE.

Although provider costs are likely to vary across the country and between hospitals, when applied to the global burden of disease and the proportions of the different pneumococcal disease syndromes, our cost estimates translate to annual provider costs of >US$110 million i.e., ∼9% of Nigeria’s 2020 health budget.[1,34] Funding of provider costs within the public sector is largely through budgetary allocations at the federal and state levels, and the overall health sector budget has been consistently below the 15% threshold of the total annual budget agreed to in the Abuja declaration. [34,35]. Treatment of pneumococcal disease exerts undue strain on the health sector, particularly at the tertiary hospitals where unit costs are higher for most components as reported elsewhere.[5,7,8] In many settings, robust pneumococcal disease surveillance has shown evidence of substantial reduction in pneumococcal diseases attributed to PCV use.[36] The extent of savings on treatment costs will depend on the effectiveness and coverage of PCV across the country.

We found substantial household costs ranging from US$44 for pneumonia to US$72 for meningitis. These figures are higher than reported for malaria in Nigeria, which ranged from ∼US$7 for outpatients to ∼US$10 for hospitalised cases;[37,38] but was lower than seen for chronic conditions, such as sickle cell anaemia (US$240) and Buruli ulcer (US$135).[39,40] These variations are attributable to differences in resource use, type of and duration of illness. User fees in our study contributed more than two-thirds of direct household costs and were largely driven by medications costs. In The Gambia, where treatment was provided at no cost to families, household OOP costs were mainly driven by non-healthcare costs including meals and visitors.[9] The main caregivers reported losing between 5-7 working days over the illness period. Although working days lost by caregivers were similar between household income levels, income loss had greater impact in the poorest households where more than half of the caregivers were also self-employed. In contrast, 80% of caregivers in the least poor households had regular salaried jobs and were likely to also receive full pay during absence for a short illness duration.

Costs incurred during treatment had considerable economic impact to households particularly for large households dependent on little income. With large numbers of economic dependents per household i.e., non-income earning household members, a one-week illness of one child, resulted in CHE_25_ in a third of households. We note the differences in CHE in ours compared to other studies in Nigeria. Although several studies analysed nationally-representative surveys, their findings differ from ours because they do not address a specific illness, target chronic conditions, and may have limited applicability to our study setting due to subnational differences in healthcare seeking behaviour.[18,41,42] At CHE_10_, the cost of treatment costs had significant burden on households regardless of their income. At higher thresholds, our results are similar to others including those that used household income as a measure of available resources like we did.[18,39,40,42,43]

Unplanned treatment expenses are likely to affect other household expenditures. OOP payment for healthcare can provide obstacles for treatment access particularly to the poorest, skewing treatment seeking towards only those than can afford to pay.[44] The higher proportion of unemployed caregivers presenting outpatient compared to inpatient may be an indicator of reduced access to hospitalised care by poorer households. The differences in household income levels between the hospitals shows a preference for the secondary hospital by poorer households. We assume that this preference is largely due to the state policy of subsidised health services for children aged <5 years in the secondary hospital.[19] Yet these households still incurred greater burden of costs as a fraction of their income, suggesting that ‘free care’ to U5s provided by the state government does not translate into lower costs to households compared to the tertiary hospital. This may be the case because drugs and medical consumables are usually excluded from these subsidised care and when they are included, they are usually out-of-stock, meaning families have to get them from other providers and often at higher OOP costs.[44] CBHI is currently being implemented in many states across Nigeria including Kano state and primarily targets the formal sector with expanding cover to vulnerable groups (women and children), informal sector and rural areas. However, we show that admissions at the state facility (MMSH) where services are supposed to ‘free’ and U5s covered by the CBHI, increased the odds of CHE_25_ almost four-fold. This suggests the benefits of the contributory scheme are yet to reach this target population, possibly due to lack of awareness of and unwillingness to pay for pre-pay for CBHI.[20]

Having a salaried job, ≥3 children U5 in the household and higher indirect costs (HCA) reduced the odds of CHE_25_. OOP payment from available household resources is the predominant way of financing healthcare in Nigeria. However, other non-health expenditures (rent, utilities and education) are financed with the same household resources as health care, and if substantial, can reduce resources ‘available’ for health expenditure.[41] We did not collect information to assess the magnitude of non-health expenses incurred by households. But having other young children or productivity losses are circumstances that can reduce resources available to households explaining their association with reduced odds of CHE. Conversely, provider costs and seeking care at a private hospital prior to hospitalisation increased odds of CHE, illustrating how burden of provider costs are pushed to households.

This study has some limitations. First, the costs here do not include provider capital costs. Second, we collected data on household costs on admission and close to or at discharge to limit bias because admission duration was short. However, there is still a risk of recall bias. Second, costs exclude children who died during admission and may have incurred costs from higher resource use due to more severe disease. Third, we only estimated time loss for the primary caregiver in hospital which does extend to fathers or other household members. Another limitation is the use of monthly income rather than household (or non-food) expenditure to assess CHE which may not identify the different ways of health financing. However, we believe current income reflects current resources and captures the current household capacity to pay for expenses of treatment given the short-term duration of illness. That majority of caregivers reported using both current income and savings to cover healthcare expenses supports our decision to use current income rather than long-term household asset. Lastly, sub-national differences in household incomes may limit generalisability of our findings, particularly the costs to household cost and its economic impact. However, some components of provider costs such as bed-day costs are not likely to differ at the tertiary hospital level because these hospitals are directly funded by the Federal Government. Additionally, because many states also offer subsidised health services to children at the secondary, our findings may be generalisable to such settings.

## CONCLUSIONS

Our analyses illustrate the treatment costs of pneumococcal disease to providers and households in Nigeria. Hospitalisation particularly at tertiary level is associated with substantial costs to both the provider and households. Households incur expenses prior to diagnosis and incur substantial direct and indirect costs that has significant impact on their incomes.

Our findings have important implications for policy. First, it is evident that the PCV programme, by averting disease, can free up scarce resources for the health sector to divert to competing health problems, reduce unexpected expenditures and CHEs, and increase resources within household for savings and essential non-health expenditures. So, it is essential to achieve and maintain high PCV coverage levels to reduce this financial burden, especially for the poorer households. Second, due to higher cost of providing care at the tertiary level, strengthening lower levels of care to provide early treatment will also significantly reduce provider costs and reduce strain on the health sector resources. Finally, the current mechanisms for financing health expenditures are inadequate to protect households from catastrophic expenses. Given that OOP payments were mainly driven by medication costs, the state government when declaring ‘free’ health services should have a realistic plan for uninterrupted supply of drugs and other essential commodities/consumables to ensure that the families are not obliged to pay for these OOP.

## Data Availability

Data supporting findings are included in the manuscript and supplement. Additional data requests can be made to the KEMRI-Wellcome Trust Research Programme Data Governance Committee.

## Acknowledgments

The authors are grateful to the staff of the paediatric units of Aminu Kano Teaching Hospital and Murtala Mohamed Specialist Hospital for facilitating the data collection.

## Ethical considerations

Written informed consent was obtained from the caregiver of each eligible child. Ethics approval for study was granted by AKTH and Kano State Ministry of Health Research Ethics Committees, the Kenya Medical Research Institute’s Scientific and Ethical Review Unit, and by the London School of Hygiene and Tropical Medicine Observational/Interventions Research Ethics Committee.

## Contributions

ALA, JAGS, IMOA conceptualised the study. ALA, IMOA, JO and JJ designed the study. ALA led the data collection with supervision from IMOA. BK led the design of database and data entry. ALA analysed the data with input from IMOA and JJ. ALA interpreted the data with guidance from IMOA, JJ, JAGS, and SG. ALA wrote the first draft. ALA, BK, MMB, MGJ, SG, JO, JAGS, JJ and IMOA reviewed successive drafts and approved the final version of the manuscript

## SUPPLEMENTARY MATERIAL

**Table S1:** Unit cost estimates and sources for parameters used in cost analysis in (US$ 2020)

| Service | Unit cost US\$ | | Source |
| --- | --- | --- | --- |
|  | AKTH | MMSH |  |
| Chest X-ray | 6.9 | 4.2 | Radiology department |
| CSF chemistry, microscopy and culture | 2.2 | 1.7 | Microbiology department |
| Blood culture | 13.9 | 3.3 | Microbiology department |
| Full blood count | 1.5 | 1.4 | Haematology department |
| Haemoglobin | 0.8 | 0.6 | Haematology department |
| Urea and electrolyte | 3.6 | 1.9 | Chemical pathology department |
| Malaria parasite | 1.1 | 0.6 | Haematology department |
| Blood transfusion | 13.9 | 13.9 | Haematology department |
| Oxygen | 20.0 | 10.0 | Paediatric ward |
| Bed day AKTH (full cost) | 55.5 | 33.3 | Paediatric ward AKTH |
| Bed day (WHO-CHOICE) | 29.8 | 16.7 | WHO-CHOICE |
| OPD visit (WHO-CHOICE) | 7.8 | 7.7 | WHO-CHOICE |

**Table S2:** Description of children and caregivers.

|  | **Outpatient**<br>**pneumonia**<br>**N=244** | **Inpatient**<br>**pneumonia**<br>**N=117** | **Septicaemia**<br>**N=66** | **Meningitis**<br>**N=53** |
| --- | --- | --- | --- | --- |
| **Hospital n (%)** |  |  |  |  |
| AKTH | 135(57.2) | 48(41.0) | 32(48.5) | 21(39.6) |
| MMSH | 109(44.9) | 69(59.0) | 34(51.5) | 32(60.4) |
| **Child characteristics** |  |  |  |  |
| **Age in months** |  |  |  |  |
| Median (IQR) | 17.9 (12-27) | 16.3 (12-26) | 24.2 (16-36) | 24.6 (18-31) |
| **Age group(months) n (%)<sup>1</sup>** |  |  |  |  |
| <1-11 | 57 (23.4) | 25 (21.4) | 6 (9.1) | 5 (9.4) |
| 12-23 | 98 (39.7) | 58 (49.6) | 26 (39.4) | 19 (35.9) |
| 24+ | 90 (36.9) | 34 (29.1) | 34 (51.5) | 29 (54.7) |
| **Gender, Female n (%)** | 119 (48.8) | 53 (45.3) | 31 (47.0) | 23 (43.4) |
| **Prior care sought n (%)** |  |  |  |  |
| None | 72 (29.5) | 25 (21.4) | 15 (22.7) | 6 (11.3) |
| Private hospital | 38 (15.6) | 17 (14.5) | 19 (28.8) | 22 (41.5) |
| Chemist | 106 (43.4) | 60 (51.3) | 28 (42.4) | 18 (34.0) |
| Others | 28 (11.5) | 15 (12.8) | 4 (6.1) | 7 (13.2) |
| Missing |  |  |  |  |
| **Caregiver characteristics** |  |  |  |  |
| Age in years, median (IQR) | 28.0 (27-30) | 29.0 (28-30) | 29.0 (28-30) | 30.0 (28-31) |
| Relationship to child,<br>Mother | 224 (91.8) | 115 (98.3) | 66 (100.0) | 49 (92.5) |
| **Highest education of caregiver n (%)** |  |  |  |  |
| None | 20 (8.2) | 7 (6.0) | 0 (0.0) | 1 (1.9) |
| Primary | 19 (7.8) | 7 (6.0) | 2 (3.0) | 3 (5.6) |
| Secondary | 88 (36.1) | 48 (41.0) | 34 (51.5) | 18 (34.0) |
| Tertiary | 117 (47.9) | 52 (44.4) | 29 (43.9) | 31 (58.5) |
| Missing | 0 (0.0) | 3 (2.6) | 1 (1.5) | 0 (0.0) |
| **Occupation of caregiver n (%)** |  |  |  |  |
| Self-employed | 98 (40.2) | 54 (46.1) | 30 (45.5) | 20 (37.7) |
| Salaried work | 97 (39.7) | 56 (47.9) | 36 (54.5) | 32 (60.4) |
| Unemployed | 49 (20.1) | 7 (6.0) | 0 (0.0) | 1 (1.9) |

**Figure S1:**
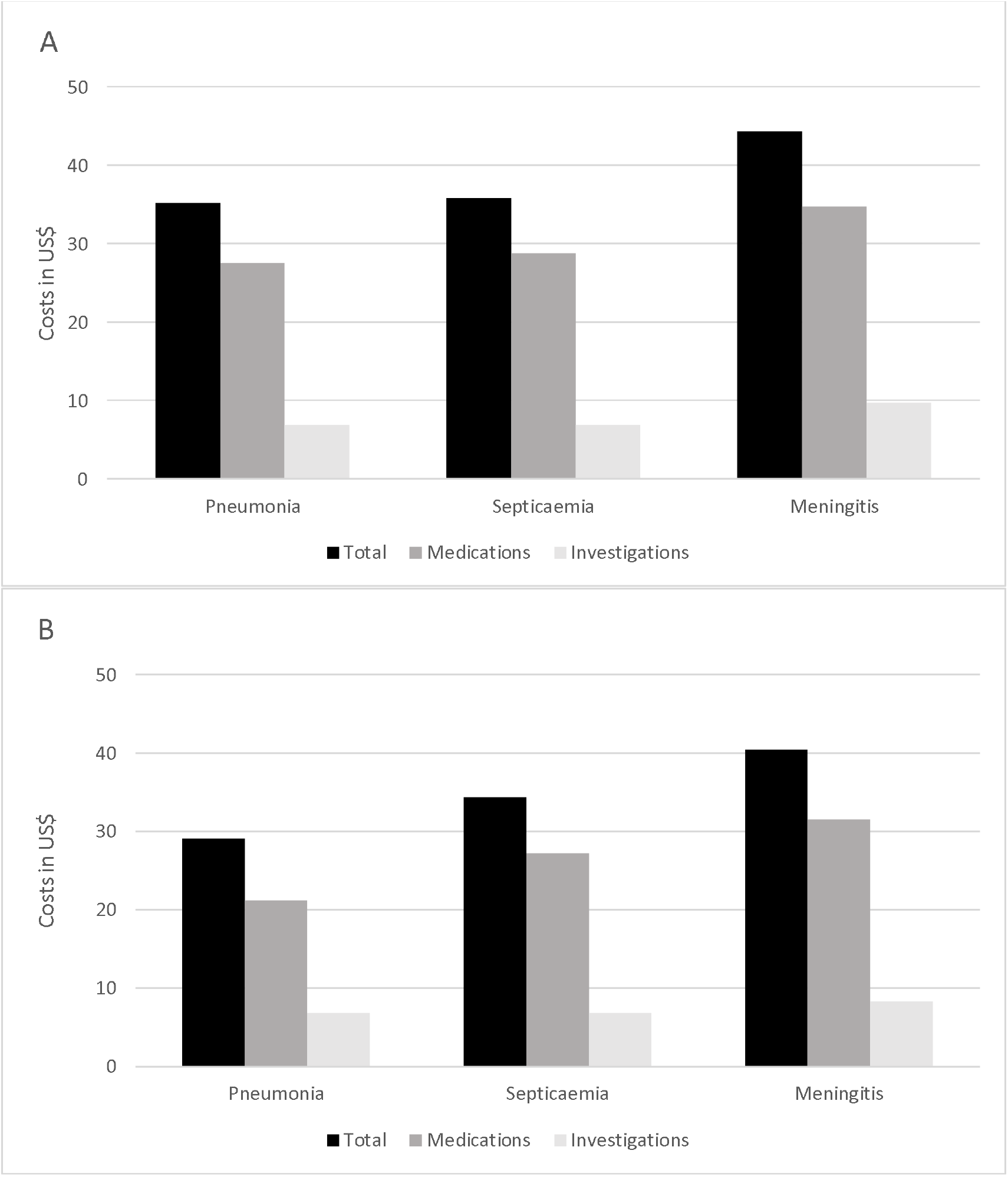
Breakdown of User fees for AKTH (A) and MMSH (B)

**Table S3:**
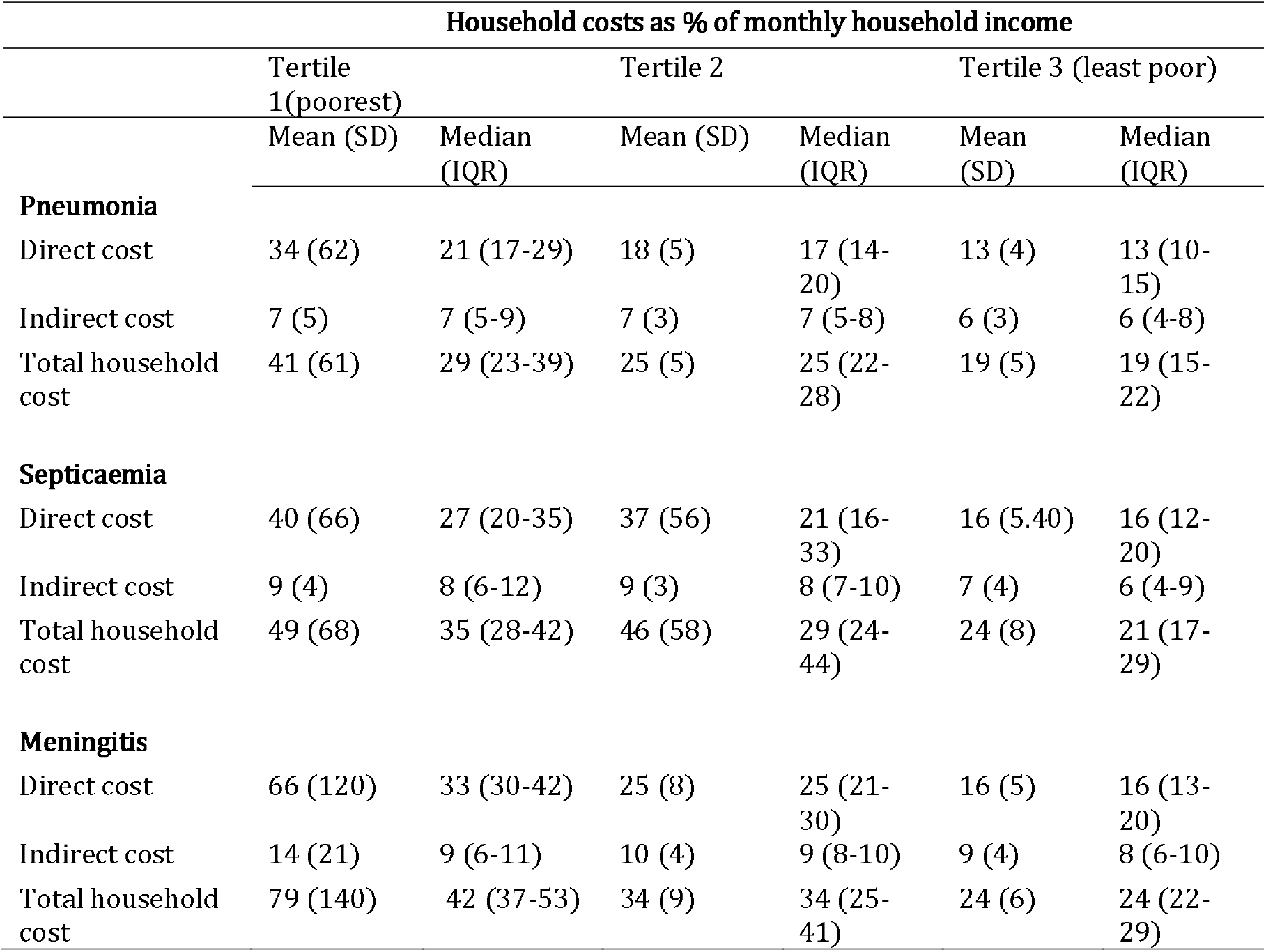
Costs incurred by households stratified by income tertiles.

|  | Household costs as % of monthly household income |  |  |  |  |  |
| --- | --- | --- | --- | --- | --- | --- |
|  | Tertile 1(poorest) |  | Tertile 2 |  | Tertile 3 (least poor) |  |
|  | Mean (SD) | Median (IQR) | Mean (SD) | Median (IQR) | Mean (SD) | Median (IQR) |
| **Pneumonia** |  |  |  |  |  |  |
| Direct cost | 34 (62) | 21 (17-29) | 18 (5) | 17 (14-20) | 13 (4) | 13 (10-15) |
| Indirect cost | 7 (5) | 7 (5-9) | 7 (3) | 7 (5-8) | 6 (3) | 6 (4-8) |
| Total household cost | 41 (61) | 29 (23-39) | 25 (5) | 25 (22-28) | 19 (5) | 19 (15-22) |
| **Septicaemia** |  |  |  |  |  |  |
| Direct cost | 40 (66) | 27 (20-35) | 37 (56) | 21 (16-33) | 16 (5.40) | 16 (12-20) |
| Indirect cost | 9 (4) | 8 (6-12) | 9 (3) | 8 (7-10) | 7 (4) | 6 (4-9) |
| Total household cost | 49 (68) | 35 (28-42) | 46 (58) | 29 (24-44) | 24 (8) | 21 (17-29) |
| **Meningitis** |  |  |  |  |  |  |
| Direct cost | 66 (120) | 33 (30-42) | 25 (8) | 25 (21-30) | 16 (5) | 16 (13-20) |
| Indirect cost | 14 (21) | 9 (6-11) | 10 (4) | 9 (8-10) | 9 (4) | 8 (6-10) |
| Total household cost | 79 (140) | 42 (37-53) | 34 (9) | 34 (25-41) | 24 (6) | 24 (22-29) |

**Table S4:** Senistivity analyses for provider costs in US$ using WHO-CHOICE estimates of bed-day costs and WTP approach for indirect costs.

| Costs US\$ | | | | | | | |
| --- | --- | --- | --- | --- | --- | --- | --- |
|  | Inpatient pneumonia |  | Septicaemia |  | Meningitis |  | P value |
|  | Mean<br>(SD) | Median (IQR) | Mean<br>(SD) | Median (IQR) | Mean<br>(SD) | Median (IQR) |  |
| **AKTH** |  |  |  |  |  |  |  |
| <u>Provider costs (WHO-CHOICE)</u> |  |  |  |  |  |  |  |
| Bed day | 146 (24) | 149 (119-149) | 159 (38) | 149 (148-179) | 189 (32) | 179 (178-208) | <0.001 |
| Total provider costs | 223 (44) | 219 (189-244) | 217 (74) | 197 (172-238) | 257 (56) | 253 (226-295) | 0.007 |
| <u>Indirect costs</u> |  |  |  |  |  |  |  |
| WTP | 25 (22) | 24 (14-31) | 41 (51) | 28 (14-53) | 56 (36) | 39 (33-83) | <0.001 |
| **MMSH** |  |  |  |  |  |  |  |
| <u>Provider costs (WHO-CHOICE)</u> |  |  |  |  |  |  |  |
| Bed day | 95 (55) | 84 (83-100) | 93 (25) | 84 (83-117) | 119 (81) | 100 (84-117) | 0.004 |
| Total provider costs | 148 (63) | 137 (120-153) | 135 (44) | 124 (108-159) | 179 (98) | 156 (131-174) | 0.001 |
| <u>Indirect costs</u> |  |  |  |  |  |  |  |
| WTP | 52 (104) | 21 (13-50) | 27 (25) | 20 (14) | 73 (134) | 33 (28-74) | 0.009 |

